# Anxiety Sensitivity as a Mediator of Internet-Based Cognitive Behavioral Therapy for Panic Disorder: A Randomized Controlled Trial with Minimal Therapist Contact

**DOI:** 10.64898/2026.05.13.26353032

**Authors:** Jorge Orrego Bravo, Rosa Raich Escursell

**Affiliations:** Universitat Autònoma de Barcelona (at time of study); atencion.org / Amindterapia.com, Barcelona, Spain; Department of Clinical and Health Psychology, Universitat Autònoma de Barcelona, Barcelona, Spain

**Keywords:** panic disorder, internet-based CBT, anxiety sensitivity, mediation, mechanism of change, minimal therapist contact, randomized controlled trial, Spain

## Abstract

**Background:** Internet-based cognitive behavioral therapy (iCBT) is efficacious for panic disorder (PD), yet the mechanisms through which it produces change remain underspecified. Anxiety sensitivity — the dispositional tendency to fear anxiety-related sensations — is theoretically central to the maintenance of PD, but its role as a mediator of iCBT outcomes has not been formally tested in Spanish-speaking populations with minimal-contact delivery formats.

**Method:** Ninety-five adults meeting DSM-IV-TR criteria for PD were randomized to an 8-week iCBT program (*Free from Anxiety*) with optional email-based therapist support (n = 49) or a waiting-list control (n = 46). Primary outcome was PD severity (PDSS); secondary outcomes included anxiety sensitivity (ASI-3), general anxiety (BAI), and depression (BDI-II). A Baron and Kenny (1986) mediation analysis with bootstrapped confidence intervals (5,000 resamples) examined ASI-3 change as a mediator of treatment effects on PDSS.

**Results:** The treatment group showed large improvements relative to controls on all outcome measures (PDSS: *d* = 0.76, 95% CI [0.10, 1.42]; mean *d* = 1.30 across measures). Mediation analysis confirmed that ASI-3 change partially mediated the treatment effect on PDSS: the indirect effect accounted for 27.4% of the total effect (indirect = 1.85; bootstrap 95% CI [0.36, 3.70]). The direct effect of treatment remained significant after controlling for ASI-3 change (*b* = 4.89, *p* < .001). Full ITT sensitivity analyses confirmed robustness (*d* = 0.47–1.47). Gains were maintained at six months (*d* = 1.15–1.26).

**Conclusions:** iCBT for PD reduces anxiety sensitivity as a partial mechanism of change, consistent with cognitive models of panic. These findings extend the evidence base for *Free from Anxiety* in Spanish clinical populations with high comorbidity and support its viability as a first-step intervention in stepped-care pathways. The anonymized dataset is publicly available at https://doi.org/10.5281/zenodo.20084725 (CC BY 4.0).

## 1. INTRODUCTION

### 1.1 From Efficacy to Mechanism

The efficacy of internet-based cognitive behavioral therapy (iCBT) for panic disorder (PD) is now well established. Meta-analyses consistently report large within-group effect sizes and outcomes broadly comparable to face-to-face CBT (Carlbring et al., 2018; Olthuis et al., 2016). Multiple randomized controlled trials have demonstrated that iCBT programs produce significant reductions in panic frequency, anticipatory anxiety, agoraphobic avoidance, and associated functional impairment (Carlbring et al., 2001, 2006; Hedman et al., 2013; Klein et al., 2006).

This accumulation of efficacy evidence has generated a productive shift in research priorities. The question is no longer *whether* iCBT works, but *how* it works — through what mechanisms, for whom, and under what conditions (Kazdin, 2007). Understanding mechanisms of change is not merely a theoretical exercise: it informs the design of more targeted interventions, identifies which program components are essential, and generates testable predictions about patient response profiles. As Kazdin (2007) noted, identifying mediators is arguably the most important methodological frontier in contemporary psychotherapy research.

### 1.2 Anxiety Sensitivity as a Candidate Mechanism

Cognitive models of PD converge on anxiety sensitivity — the fear of anxiety-related sensations

— as a core maintaining mechanism (Clark, 1986; Barlow, 2002). According to Clark’s (1986) model, panic attacks are sustained by a catastrophic misinterpretation cycle: benign physiological sensations are perceived as dangerous, amplifying anxiety, intensifying sensations, and apparently confirming the perceived threat. Anxiety sensitivity indexes the dispositional tendency to engage in this cycle, and its reduction is theoretically necessary for durable symptom improvement.

Empirically, elevated anxiety sensitivity is consistently observed in clinical PD populations (Taylor et al., 2007), and its reduction through CBT mediates panic symptom improvement in face-to-face treatment (Smits et al., 2004; Bouchard et al., 2007). However, formal mediation tests in iCBT contexts are rare — and none, to our knowledge, have been conducted in Spanish-speaking populations or with minimal-contact delivery formats. This gap is meaningful because the relative absence of relational scaffolding in minimal-contact iCBT may concentrate therapeutic action more directly on cognitive-behavioral content, potentially rendering anxiety sensitivity reduction a more prominent pathway than in therapist-intensive formats where alliance and motivational processes also contribute.

## 1.3 The Present Study

The present trial evaluates the *Free from Anxiety* iCBT program delivered with minimal optional email contact versus a waiting-list control in a Spanish adult clinical population. The primary aim is to test whether reduction in anxiety sensitivity (ASI-3) mediates the treatment effect on panic severity (PDSS) using bootstrapped mediation analysis. Secondary aims characterize the efficacy profile of this minimal-contact format — including sensitivity ITT analyses, six-month maintenance, and educational level as a potential moderator — in a sample with high clinical comorbidity representative of real-world presentations.

We hypothesized that: (1) the treatment group would show significantly greater PDSS improvement than controls; (2) ASI-3 change would partially mediate this effect, with the bootstrap CI for the indirect effect excluding zero; and (3) a significant direct treatment effect would remain after controlling for ASI-3, reflecting additional behavioral and pharmacological mechanisms.

## 2. METHOD

### 2.1 Participants

Participants were recruited through targeted online advertisement and directed to a study website. Eligibility criteria required: (a) DSM-IV-TR diagnosis of PD with or without agoraphobia, confirmed by structured clinical interview (MINI); (b) PD duration ≥ 12 months; (c) PD as primary presenting pathology; (d) stable medication dosage for ≥ 3 months if currently medicated; (e) age 18–60; and (f) Spanish language competency and residency in Spain. Exclusion criteria comprised: (a) comorbid disorder requiring immediate clinical attention; (b) BDI-II score > 25; and (c) current or prior CBT for PD.

Of 274 individuals who expressed initial interest, 152 completed online screening. Of these, 95 met eligibility criteria and were randomized. Excluded candidates (n = 57) were referred to the public healthcare system with individualized feedback on their screening results. The complete participant flow is presented in Figure 1 (CONSORT diagram). The study received ethical approval from the Animal and Human Experimentation Ethics Committee of the Universitat Autònoma de Barcelona (approval date: 26 July 2013; ClinicalTrials.gov NCT02402322). Written informed consent was obtained from all participants prior to inclusion.

**Figure 1.**
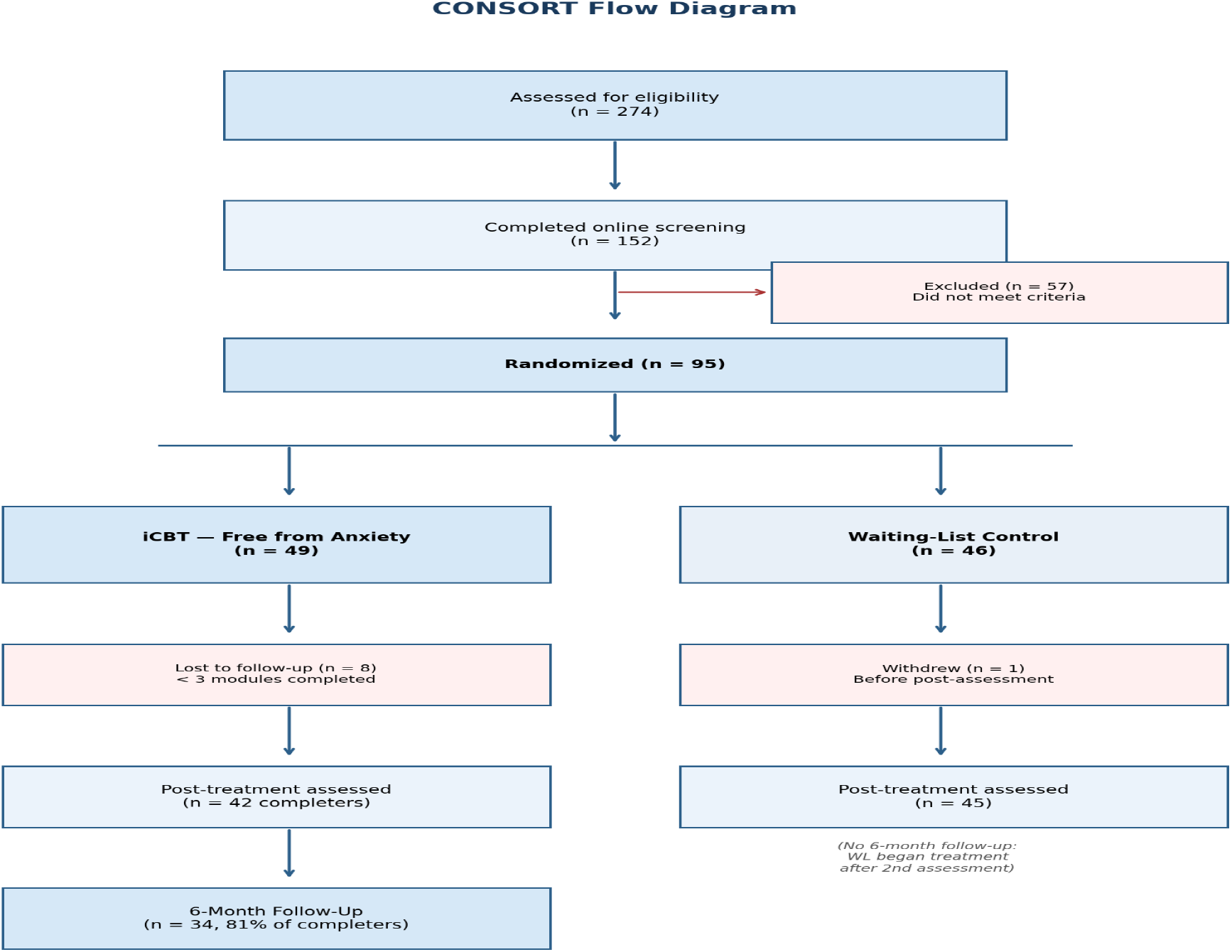
CONSORT flow diagram of participant enrollment, allocation, follow-up, and analysis.

**Figure 2.**
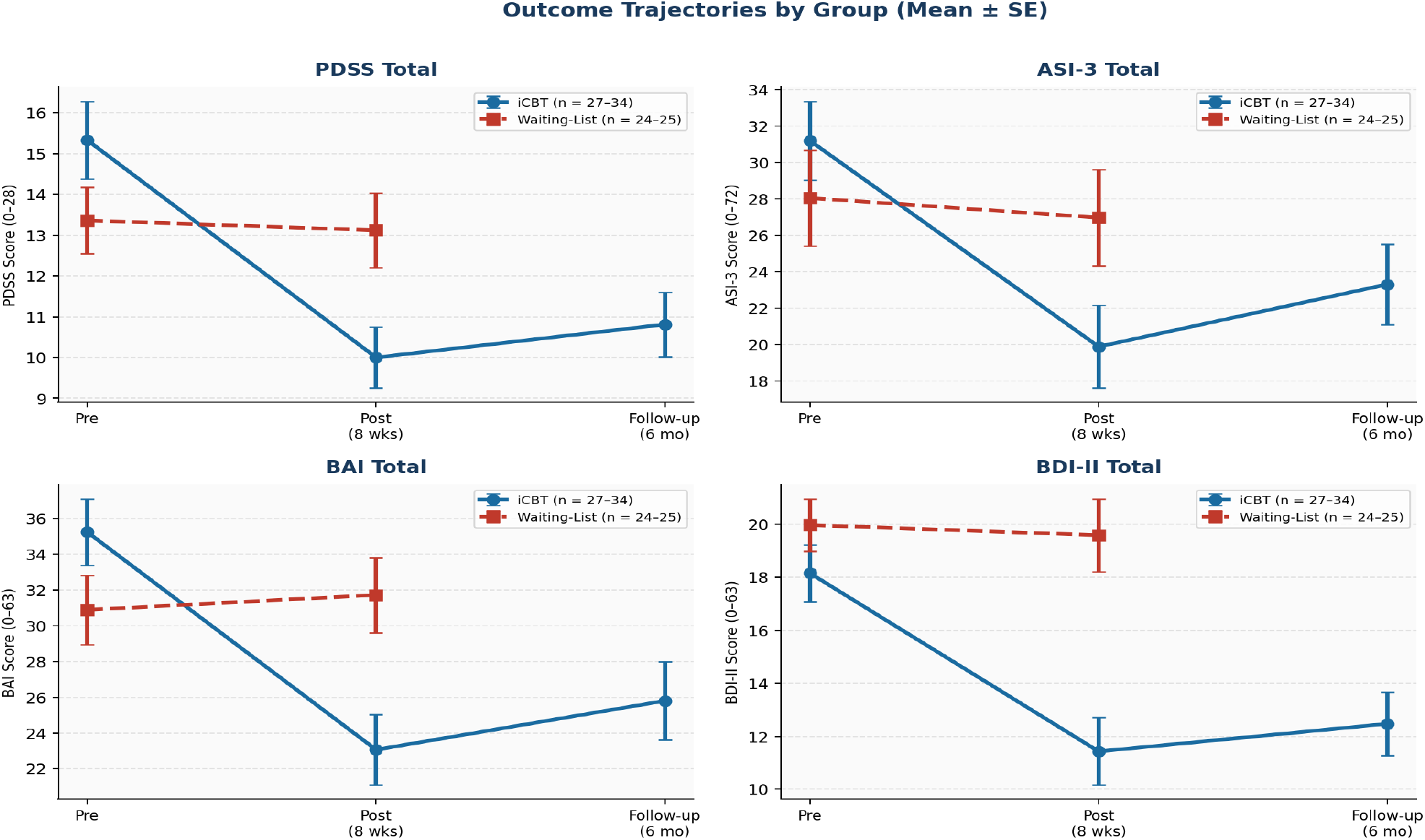
Mean outcome scores (±1 SE) by group across assessment points. WL = waiting-list.

### 2.2 Measures

#### PDSS (primary outcome)

The Panic Disorder Severity Scale (Shear et al., 1992; Spanish: Santacana et al., 2014; α = .85) is a 7-item scale assessing PD severity across panic frequency, distress, anticipatory anxiety, agoraphobic and interoceptive avoidance, and functional impairment (range 0–28; ≥ 8 clinically significant; ≥ 16 severe).

#### ASI-3 (hypothesized mediator)

The Anxiety Sensitivity Index-3 (Taylor et al., 2007; Spanish: Sandín & Valiente, 2007; α = .91) is an 18-item self-report scale measuring the dispositional tendency to fear anxiety-related sensations across physical, cognitive, and social dimensions (range 0–72).

#### BAI

The Beck Anxiety Inventory (Beck et al., 1988; Spanish: Sanz & Navarro, 2003; α = .93) is a 21-item self-report measure of anxiety symptom severity (range 0–63).

#### BDI-II. The Beck Depression Inventory-II (Beck et al., 1996; Spanish: Sanz & Vázquez, 1998; α = .89) is a 21-item self-report measure of depressive symptom severity (range 0–63)

#### SDI

The Sheehan Disability Inventory (Sheehan et al., 1996; Spanish: Bobes et al., 1999; α = .72) is a 5-item instrument assessing functional impairment in work, social, and family life domains (range 0–10 per subscale).

### 2.3 Intervention: Free from Anxiety

*Free from Anxiety* (*Fri från oro*) is a transdiagnostic iCBT program originally developed in Sweden by Ström et al. (2000), adapted to Spanish by the first author under license from Livanda (Sweden). The program consists of 8 weekly interactive modules delivered via a secure online platform, covering: psychoeducation (all modules), behavioral analysis (modules 1–2), interoceptive and in-vivo exposure (modules 2–8), progressive relaxation (modules 1–4), cognitive restructuring (modules 3–5), mindfulness (module 6), communication skills (module 7), and problem-solving (module 8). The first three modules contain PD-specific content and constitute the minimum threshold for meaningful intervention exposure.

### 2.4 Design and Procedure

A two-arm parallel-group RCT was employed. Following diagnostic assessment, eligible participants were randomized to treatment (n = 49) or waiting-list control (n = 46) via computer-generated randomization by an independent researcher.

#### Treatment condition

Participants accessed the iCBT program independently and could contact their assigned psychologist via the platform’s integrated email system at any time to resolve doubts about technical or content-related matters. Psychologists responded within 24 hours. No proactive outreach was initiated by clinicians. Given the nature of the intervention, therapist blinding was not feasible; all outcome measures were self-reported, minimizing assessor bias.

#### Waiting-list condition

Participants received no active treatment during the 8-week period. Following post-treatment assessment, they were offered access to the iCBT program. As the waiting-list group began treatment immediately after the second assessment, they were not followed up at six months. Assessments were conducted at pre-treatment (baseline), post-treatment (week 8), and six-month follow-up (treatment group only).

### 2.5 Statistical Analyses

Baseline group equivalence was examined with independent-samples *t*-tests and chi-square tests. Primary between-group comparisons used two-way repeated-measures ANOVA (time × group) followed by Bonferroni-corrected post-hoc *t*-tests (corrected α = .017). Effect sizes were computed as Cohen’s *d* with pooled standard deviations; 95% CIs used the noncentral *t* approximation.

#### Mediation analysis

A Baron and Kenny (1986) mediation model was estimated with treatment condition (1 = treatment, 0 = WL) as the independent variable (X), pre-to-post change in ASI-3 as the mediator (M), and pre-to-post change in PDSS as the outcome (Y), in the complete post-treatment sample (n = 73 with full data). The indirect effect (a × b) was tested via the Sobel test and confirmed with 5,000 bootstrap resamples; a 95% bootstrap CI excluding zero was taken as evidence of mediation. The proportion of total effect mediated was computed as (total − direct) / total.

#### Sensitivity analyses

A full intention-to-treat (ITT) analysis was conducted with missing post-treatment values imputed using last observation carried forward (LOCF). Post-hoc power analyses were conducted for all outcome comparisons. Educational level (low/medium/high) was tested as a moderator via ANOVA on post-treatment scores. All analyses were conducted in SPSS v.22.

## 3. RESULTS

### 3.1 Sample Characteristics and Baseline Equivalence

The 95 randomized participants had a mean age of 40.46 years (*SD* = 9.83, range 20–60); 63.2% were female. The sample presented high clinical complexity: 48.1% met criteria for comorbid depression and 25.3% for comorbid social phobia — rates substantially higher than those typical of RCT samples and more representative of real-world clinical presentations. No significant baseline differences emerged between groups on any demographic variable or pre-treatment outcome measure (all *p* > .05; see Table 1). Eight treatment participants completed fewer than three modules and were classified as dropouts; one control participant withdrew before post-treatment assessment. Post-treatment data were available for 87 participants (91.6%). Only 8 of 49 treatment participants used the available email support at any point; contacts were brief and procedural. The program thus functioned as an effectively unguided self-help intervention for the large majority of participants.

**Table 1.**
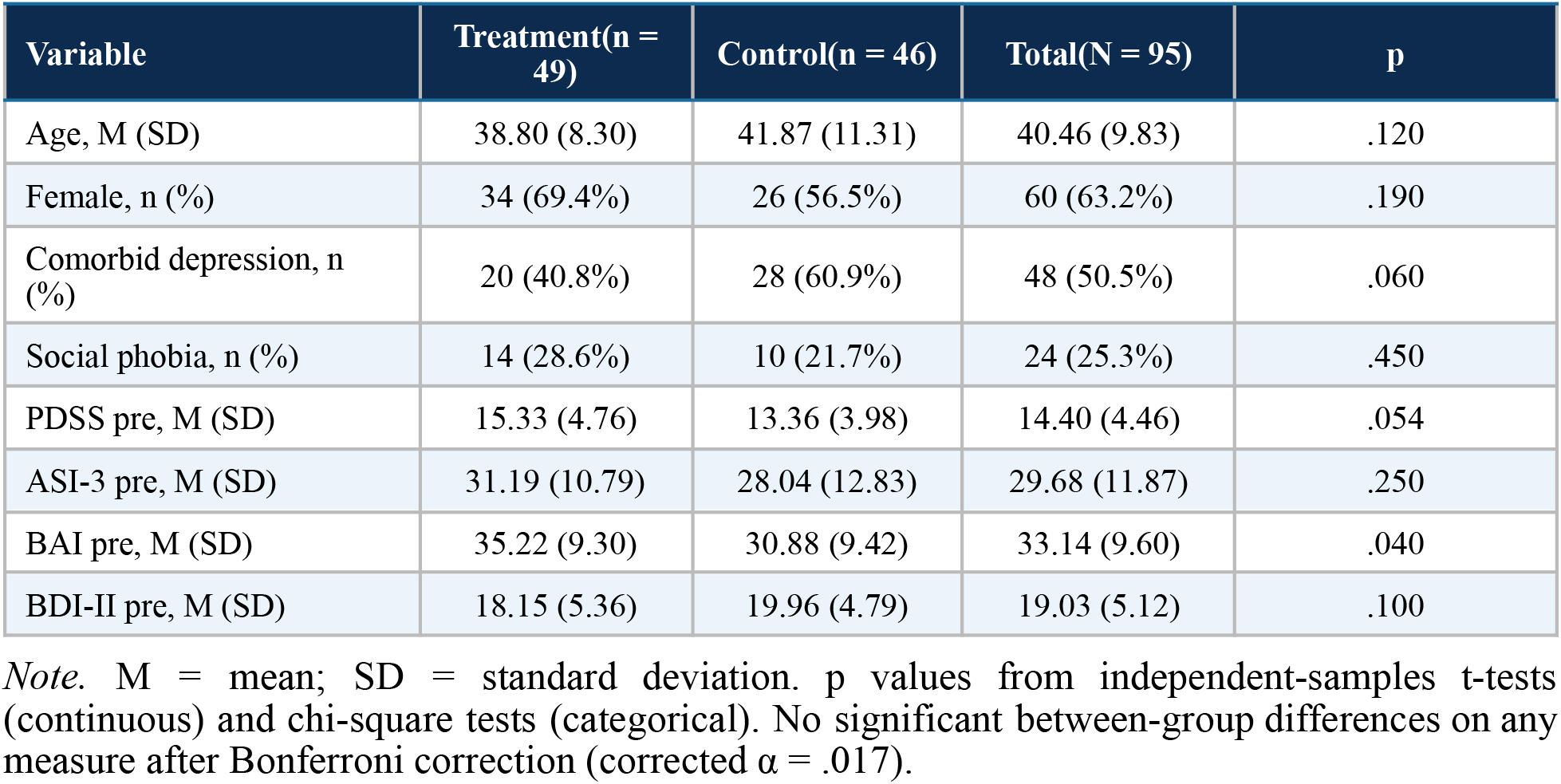
Baseline Demographic and Clinical Characteristics by Group.

| Variable | Treatment(n = 49) | Control(n = 46) | Total(N = 95) | p |
| --- | --- | --- | --- | --- |
| Age, M (SD) | 38.80 (8.30) | 41.87 (11.31) | 40.46 (9.83) | .120 |
| Female, n (%) | 34 (69.4%) | 26 (56.5%) | 60 (63.2%) | .190 |
| Comorbid depression, n (%) | 20 (40.8%) | 28 (60.9%) | 48 (50.5%) | .060 |
| Social phobia, n (%) | 14 (28.6%) | 10 (21.7%) | 24 (25.3%) | .450 |
| PDSS pre, M (SD) | 15.33 (4.76) | 13.36 (3.98) | 14.40 (4.46) | .054 |
| ASI-3 pre, M (SD) | 31.19 (10.79) | 28.04 (12.83) | 29.68 (11.87) | .250 |
| BAI pre, M (SD) | 35.22 (9.30) | 30.88 (9.42) | 33.14 (9.60) | .040 |
| BDI-II pre, M (SD) | 18.15 (5.36) | 19.96 (4.79) | 19.03 (5.12) | .100 |

*Note.* M = mean; SD = standard deviation. p values from independent-samples t-tests (continuous) and chi-square tests (categorical). No significant between-group differences on any measure after Bonferroni correction (corrected $\alpha = .017$ ).

### 3.2 Post-Hoc Power Analysis

Post-hoc power analyses (*n*treatment = 42, *n*control = 45, α = .05, two-tailed) indicated adequate power for the primary and most secondary outcomes: power exceeded 97% for the PDSS (*d* = 1.47), BAI (*d* = 0.86), and BDI-II (*d* = 1.25). The minimum detectable effect at 80% power was *d* = 0.61. The ASI-3 between-group comparison (*d* = 0.58) fell marginally below this threshold (power = 76.2%), indicating a slightly elevated Type II error risk for that specific comparison. A formal a priori power calculation was not conducted; this is acknowledged as a limitation.

### 3.3 Treatment Efficacy: Primary and Secondary Outcomes

Repeated-measures ANOVA revealed significant Time × Group interactions on all four primary measures (all *p* < .001). Full descriptive statistics, within-group effect sizes, and between-group comparisons are presented in Tables 2 and 3.

**Table 2.** Pre-, Post-Treatment, and Follow-Up Scores by Group with Within-Group Effect Sizes.

| Measure | Group | n | Pre M (SD) | Post M (SD) | Follow-up M (SD) | d [95% CI] |
| --- | --- | --- | --- | --- | --- | --- |
| **PDSS** | Tx | 25–27 | 15.33 (4.76) | 10.00 (3.72) | 10.81 (3.98) | 1.47 [0.84, 2.09] |
|  | WL | 24–25 | 13.36 (3.98) | 13.12 (4.50) | — | –0.02 [–.58, .54] |
| **ASI-3** | Tx | 26–27 | 31.19 (10.79) | 19.88 (11.30) | 23.30 (11.04) | 1.32 [0.72, 1.92] |
|  | WL | 25 | 28.04 (12.83) | 26.96 (12.97) | — | 0.08 [–.48, .63] |
| **BAI** | Tx | 25–27 | 35.22 (9.30) | 23.08 (9.90) | 25.81 (10.88) | 1.88 [1.22, 2.55] |
|  | WL | 24–25 | 30.88 (9.42) | 31.71 (10.27) | — | –0.16 [–.72, .40] |
| **BDI-II** | Tx | 25–27 | 18.15 (5.36) | 11.44 (6.36) | 12.48 (5.94) | 1.41 [0.79, 2.03] |
|  | WL | 24–25 | 19.96 (4.79) | 19.58 (6.72) | — | 0.03 [–.53, .59] |

*Note.* Tx = treatment group; WL = waiting-list group; d = within-group Cohen's d (pre vs. post); 95% CI for d. Treatment group: all $p < .001$ . WL group: all $p > .40$ . WL not assessed at follow-up. PDSS = Panic Disorder Severity Scale (range 0–28); ASI-3 = Anxiety Sensitivity Index-3 (range 0–72); BAI = Beck Anxiety Inventory (range 0–63); BDI-II = Beck Depression Inventory-II (range 0–63).

**Table 3.** Between-Group Post-Treatment Comparisons (mITT) and Full ITT Sensitivity Effect Sizes.

| Measure | M diff | 95% CI | t | p | d | 95% CI (d) | ITT d |
| --- | --- | --- | --- | --- | --- | --- | --- |
| PDSS | 3.12 | [0.82, 5.43] | 2.65 | .011 | **0.76** | [0.10, 1.42] | 0.56 |
| ASI-3† | 7.08 | [0.41, 13.74] | 2.08 | .043 | 0.58 | [-0.07, 1.23] | 0.47 |
| BAI | 8.63 | [2.98, 14.28] | 2.99 | .004 | **0.86** | [0.20, 1.52] | 0.65 |
| BDI-II | 8.14 | [4.48, 11.81] | 4.36 | < .001 | **1.25** | [0.57, 1.93] | 1.10 |

*Note.* M diff = WL minus Tx (positive values indicate WL scored higher). Bonferroni-corrected $\alpha = .017$ . ITT d = effect size under full ITT analysis with LOCF imputation. Bold d values exceed the conventional large-effect threshold ( $d \geq 0.80$ ). † $p < .05$ but above Bonferroni threshold; treated as marginal.

For the primary outcome, the treatment group demonstrated a mean PDSS reduction of 33.0% from pre-to post-treatment (pre: *M* = 15.33, *SD* = 4.76; post: *M* = 10.00, *SD* = 3.72; *d* = 1.47, 95% CI [0.84, 2.09]), compared with no significant change in the waiting-list group (pre: *M* = 13.36, *SD* = 3.98; post: *M* = 13.12, *SD* = 4.50; *d* = −0.02). The between-group effect at post-treatment was large (*d* = 0.76, 95% CI [0.10, 1.42]; mean difference = 3.12, 95% CI [0.82, 5.43], *p* = .011). Between-group effect sizes across secondary outcomes ranged from *d* = 0.58 (ASI-3) to *d* = 1.25 (BDI-II). The waiting-list group showed no significant change on any measure (all *p* > .40).

### 3.4 Primary Finding: Mediation by Anxiety Sensitivity

All four Baron and Kenny (1986) conditions for mediation were satisfied (n = 73 with complete data). Results are summarized below and illustrated in Figure 3.

**Figure 3.**
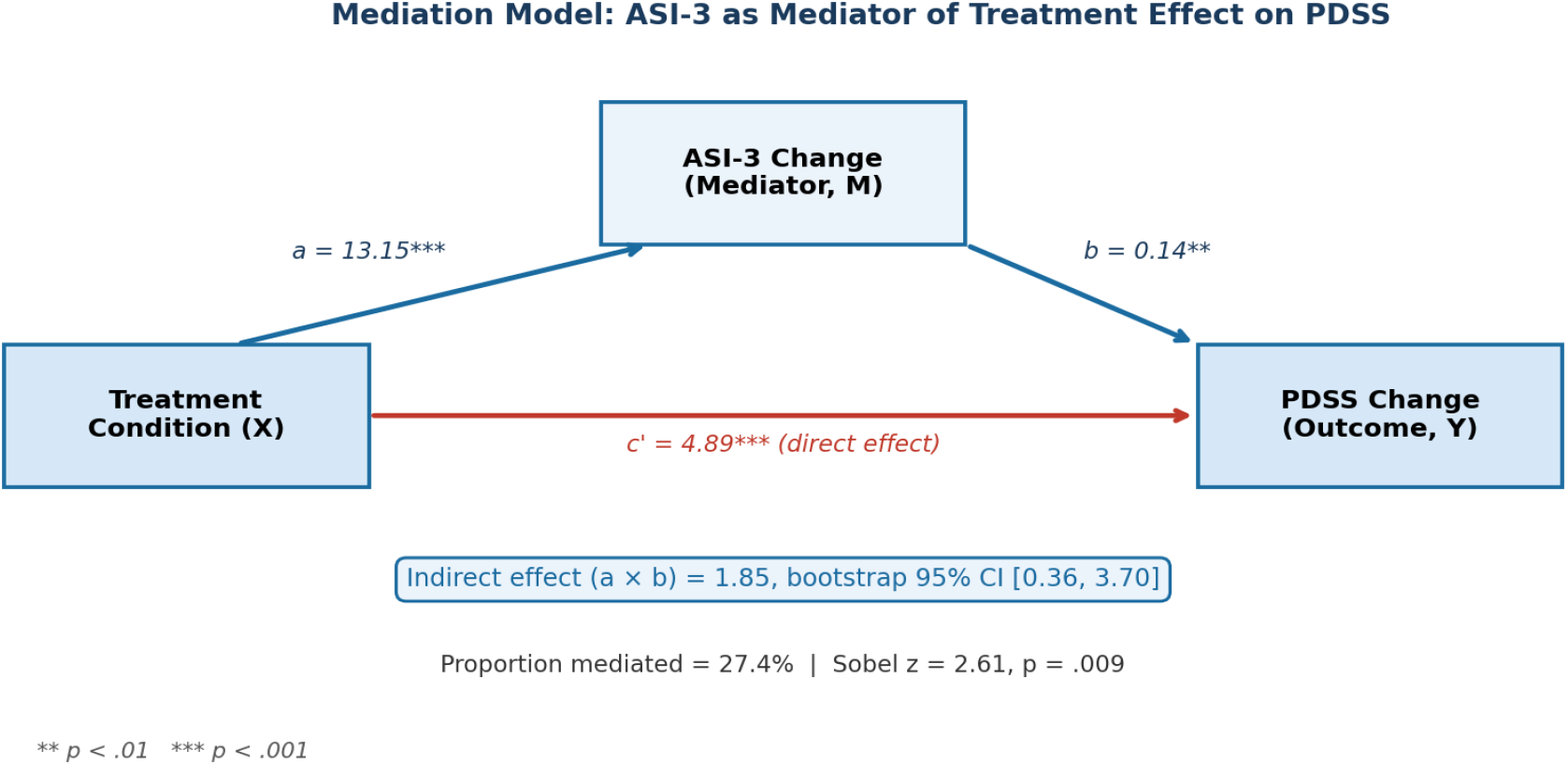
Baron and Kenny (1986) mediation model. Path coefficients are unstandardized. ** p < .01, *** p < .001.

**Figure 4.**
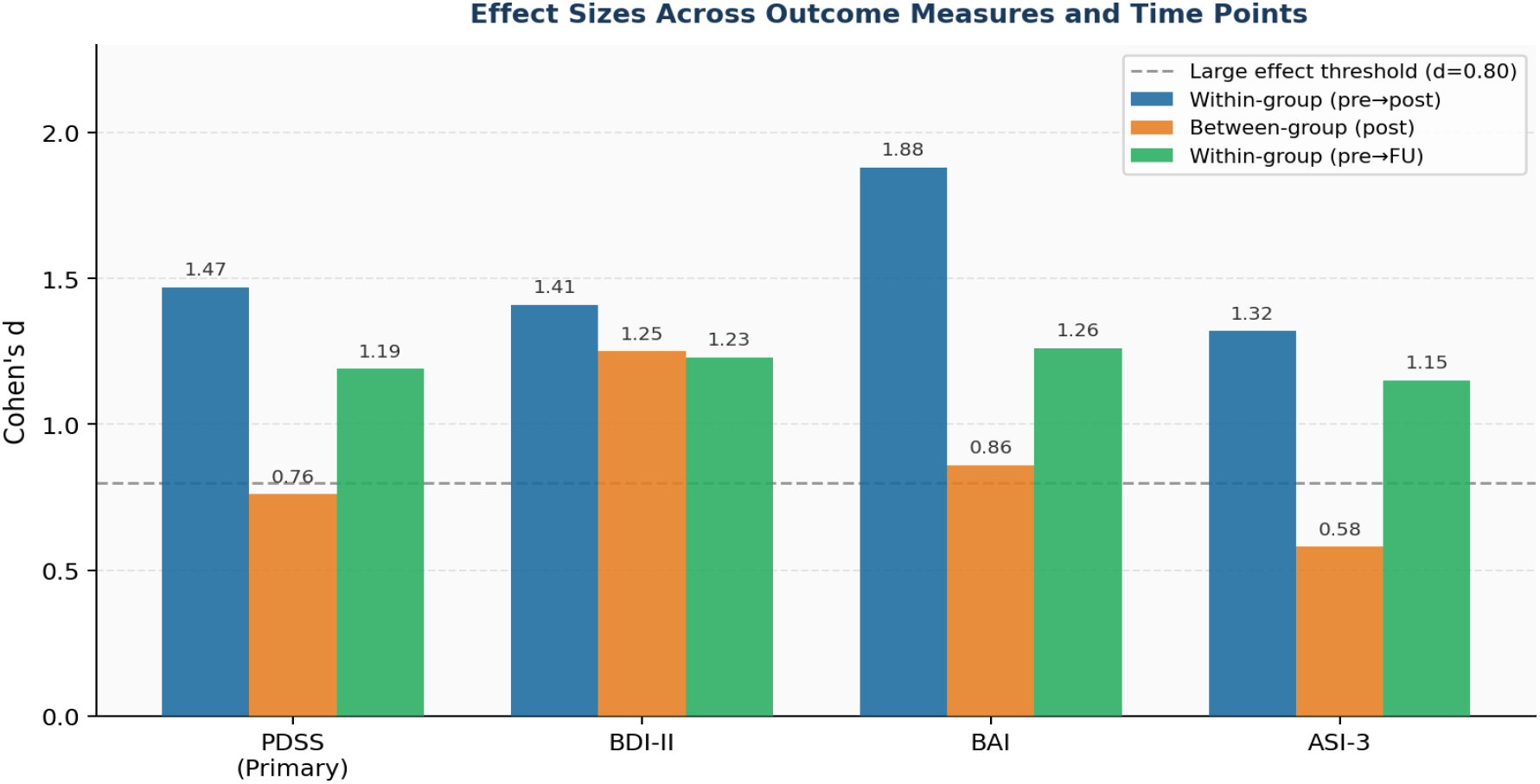
Cohen’s d effect sizes across outcome measures and time points. Dashed line indicates the conventional large-effect threshold (d = 0.80).

**Step 1 (c path)**. Treatment significantly predicted PDSS change: *b* = 6.74, *SE* = 0.87, *t*(71) = 7.73, *p* < .001.

**Step 2 (a path)**. Treatment significantly predicted ASI-3 change: *b* = 13.15, *SE* = 1.99, *t*(71) = 6.60, *p* < .001.

**Step 3 (b path)**. ASI-3 change significantly predicted PDSS change controlling for treatment: *b* = 0.14, *SE* = 0.05, *t*(70) = 2.84, *p* = .006.

**Step 4 (c’ path)**. The direct treatment effect on PDSS remained significant after controlling for ASI-3: *b* = 4.89, *SE* = 1.06, *t*(70) = 4.63, *p* < .001.

The Sobel test confirmed the indirect effect: indirect = 1.85, *SE* = 0.71, *z* = 2.61, *p* = .009. Bootstrap analysis (5,000 resamples) produced a 95% CI of [0.36, 3.70], which excludes zero. The proportion of the total treatment effect mediated by ASI-3 change was 27.4%, indicating partial mediation.

### 3.5 Sensitivity Analysis: Full ITT with LOCF

Full ITT analyses (14 values imputed via LOCF) yielded uniformly smaller effect sizes (Δ*d* = −0.03 to −0.26) relative to the primary mITT analysis, confirming that mITT estimates were slightly inflated. Despite this attenuation, all primary between-group comparisons remained statistically significant and large (*d* = 0.47–1.47; all *p* < .05 except ASI-3, *p* = .095). The qualitative pattern of findings was unchanged (see Table 3, ITT *d* column).

### 3.6 Six-Month Follow-Up

Treatment gains were fully maintained at six months. Within-group pre-to-follow-up effect sizes ranged from *d* = 1.15 (ASI-3) to *d* = 1.26 (BAI). No significant deterioration occurred from post-treatment to follow-up on any measure (all *p* > .05). PDSS scores showed a non-significant trend toward further improvement beyond program completion (post: *M* = 10.00; follow-up: *M* = 10.81).

### 3.7 Educational Level as Moderator

Educational level (low/medium/high) did not moderate treatment outcomes. Pre-treatment PDSS scores did not differ across educational groups (*p* = .865), nor did post-treatment scores (*p* = .563), indicating that the program produced comparable effects regardless of formal educational attainment.

## 4. DISCUSSION

### 4.1 Anxiety Sensitivity as a Mechanism of Change

The central finding of this trial is that reduction in anxiety sensitivity partially mediates the effect of iCBT on panic severity, with a bootstrap-confirmed indirect effect accounting for 27.4% of the total treatment effect. This is, to our knowledge, the first formal demonstration of this mediation pathway in a Spanish clinical population receiving iCBT, and the first to test it under minimal-contact delivery conditions.

The finding is theoretically significant for several reasons. First, it provides direct empirical support for Clark’s (1986) cognitive model of panic in the context of internet-delivered treatment: the program’s psychoeducational and exposure components appear to modify the specific cognitive-behavioral mechanism the model identifies as central — the tendency to catastrophically interpret somatic anxiety signals. Second, the partial nature of the mediation is informative: the direct treatment effect remained large and significant (*b* = 4.89, 72.6% of total effect), indicating that anxiety sensitivity reduction is one of multiple active pathways. Behavioral mechanisms (reduction in agoraphobic avoidance via in-vivo exposure) and pharmacological effects in medicated participants likely account for the residual direct effect.

Third, the finding has implications for program design. If anxiety sensitivity reduction is a key mechanism, then the modules most directly targeting it — interoceptive exposure and cognitive restructuring of somatic misinterpretations (modules 2–5) — are not optional enhancements but core active ingredients. Early assessment of ASI-3 change could function as a within-treatment response indicator to guide stepped-care decisions.

### 4.2 Efficacy Under Real-World Clinical Complexity

The efficacy findings extend the evidence base for *Free from Anxiety* in a clinical population with substantially higher comorbidity than typical RCT samples. With 48.1% comorbid depression and 25.3% social phobia, this sample more closely approximates the presentations encountered in routine care. The large between-group effect sizes (*d* = 0.58–1.25 under mITT; *d* = 0.47–1.10 under full ITT), maintained across six months, indicate that the program is robustly efficacious under these conditions.

Notably, the program functioned as an effectively unguided intervention — fewer than 17% of participants used the available email support — yet produced effects substantially exceeding the meta-analytic benchmark for CBT-based PD treatments of *d* = 1.19 (Sánchez-Meca et al., 2010). This positions minimal-contact iCBT as a viable first-step intervention in stepped-care frameworks, with structured weekly guidance (Oromendia et al., 2016) available as an evidence-based escalation option for non-responders.

### 4.3 Equity of Access: Educational Level as Non-Moderator

The absence of educational moderation (all *p* > .56) has direct implications for deployment equity. A recurrent concern in the iCBT literature is that internet-based interventions may systematically favor more educated, digitally literate users, reproducing existing health inequalities (Christensen et al., 2009; Marks et al., 2007). The present data do not support this concern for *Free from Anxiety* in a Spanish clinical population: the program produced comparable effects across all educational levels, supporting its potential for equitable deployment in public health pathways.

### 4.4 Limitations

Several limitations require acknowledgment. (1) A formal a priori power calculation was not conducted; post-hoc analyses indicate adequate power for most comparisons (> 97%) but the ASI-3 between-group comparison was marginally underpowered (76.2%). (2) mITT analyses may have slightly inflated effect sizes; full ITT analyses confirm qualitative robustness. (3) Therapist blinding was not feasible; all outcome measures were self-reported. (4) The sample was self-referred via online advertisement, limiting generalizability to populations less proactively seeking digital treatment. (5) Six-month follow-up was available only for the treatment group. (6) While the mediation analysis satisfies Baron and Kenny’s (1986) conditions and produces a bootstrap-confirmed indirect effect, the cross-sectional nature of the pre-to-post data precludes causal inference about the temporal ordering of ASI-3 and PDSS change within the treatment period. Future research should incorporate within-treatment assessment points to test the temporal mediation hypothesis directly.

## 5. CONCLUSIONS

This trial provides three integrated contributions to the iCBT literature for panic disorder. First, it demonstrates large and durable treatment efficacy under minimal-contact conditions in a Spanish clinical population with high real-world comorbidity, with effects robust to full ITT sensitivity analyses and maintained at six months. Second, it provides the first bootstrap-confirmed mediation analysis demonstrating that anxiety sensitivity reduction is a partial mechanism of treatment action, accounting for 27.4% of the total effect on panic severity — consistent with cognitive models of PD and with direct implications for program design and within-treatment monitoring. Third, educational level does not moderate outcomes, supporting the program’s potential for equitable deployment across heterogeneous populations.

Together, these findings position *Free from Anxiety* as an efficacious, mechanistically grounded, and potentially equitable first-step intervention for PD in Spanish-speaking health systems. The publicly available anonymized dataset (https://doi.org/10.5281/zenodo.20084725; CC BY 4.0) enables independent replication, meta-analytic inclusion, and secondary research.

## Data Availability

All data generated in this study are available from the authors upon reasonable request

https://doi.org/10.5281/zenodo.20084725

## DECLARATIONS

### Ethics Statement

The Animal and Human Experimentation Ethics Committee of the Universitat Autònoma de Barcelona granted ethical approval for this work (approval date: 26 July 2013). The study was prospectively registered at ClinicalTrials.gov (NCT02402322) prior to participant recruitment.

### Informed Consent Statement

Written informed consent was obtained from all participants prior to their inclusion in the study. Participants were informed of their right to withdraw at any time without consequence. All personal data were treated confidentially and used exclusively for academic and research purposes.

### Data Availability

The anonymized dataset supporting the findings of this study is publicly available at Zenodo under a Creative Commons Attribution 4.0 International (CC BY 4.0) license: https://doi.org/10.5281/zenodo.20084725. The dataset includes all outcome variables across pre-treatment, post-treatment, and six-month follow-up assessment points, accompanied by a full codebook.

### Author Contributions

J.O.B. conceptualized the study, developed and adapted the intervention, conducted data collection, performed statistical analyses, and wrote the original manuscript. R.R.E. supervised the study design and data collection and critically reviewed and edited the manuscript. Both authors approved the final version.

## REFERENCES

Barlow, D. H. (2002). Anxiety and its disorders: The nature and treatment of anxiety and panic (2nd ed.). Guilford Press.

Baron, R. M., & Kenny, D. A. (1986). The moderator-mediator variable distinction in social psychological research. Journal of Personality and Social Psychology, 51(6), 1173–1182. 10.1037/0022-3514.51.6.1173

Beck, A. T., Epstein, N., Brown, G., & Steer, R. A. (1988). An inventory for measuring clinical anxiety. Journal of Consulting and Clinical Psychology, 56, 893–897.

Beck, A. T., Steer, R. A., Ball, R., & Ranieri, W. F. (1996). Comparison of Beck Depression Inventories–IA and –II in psychiatric outpatients. Journal of Personality Assessment, 67, 588–597.

Bobes, J. (1998). A Spanish validation study of the Mini International Neuropsychiatric Interview. European Psychiatry, 13, 198s–199s.

Bobes, J., Badía, X., Luque, A., García, M., González, M. P., & Dal-Ré, R. (1999). Validación de las versiones en español del SDI. Medicina Clínica, 112, 530–538.

Bouchard, S., Gauthier, J., Nouwen, A., Ivers, H., Vallières, A., Simard, S., & Fournier, T. (2007). Temporal relationship between dysfunctional beliefs, self-efficacy and panic apprehension in the treatment of panic disorder with agoraphobia. Journal of Behavior Therapy and Experimental Psychiatry, 38(3), 275–292.

Carlbring, P., Andersson, G., Cuijpers, P., Riper, H., & Hedman-Lagerlöf, E. (2018). Internet-based vs. face-to-face cognitive behavior therapy for psychiatric and somatic disorders: An updated systematic review and meta-analysis. Cognitive Behaviour Therapy, 47(1), 1–18. 10.1080/16506073.2017.1401115

Carlbring, P., Bohman, S., Brunt, S., Buhrman, M., Westling, B. E., Ekselius, L., & Andersson, G. (2006). Remote treatment of panic disorder: A randomized trial of internet-based CBT supplemented with telephone calls. American Journal of Psychiatry, 163, 2119–2125.

Carlbring, P., Westling, B. E., Ljungstrand, P., Ekselius, L., & Andersson, G. (2001). Treatment of panic disorder via the internet: A randomized trial of a self-help program. Behavior Therapy, 32, 751–764.

Christensen, H., Griffiths, K. M., & Farrer, L. (2009). Adherence in internet interventions for anxiety and depression. Journal of Medical Internet Research, 11, e13. 10.2196/jmir.1194

Clark, D. M. (1986). A cognitive approach to panic. Behaviour Research and Therapy, 24(4), 461–470. 10.1016/0005-7967(86)90116-9

Hedman, E., Ljótsson, B., Rück, C., Bergström, J., Andersson, G., Kaldo, V., & Lindefors, N. (2013). Effectiveness of internet-based CBT for panic disorder in routine psychiatric care. Acta Psychiatrica Scandinavica, 128, 457–467. 10.1111/acps.12079

Kazdin, A. E. (2007). Mediators and mechanisms of change in psychotherapy research. Annual Review of Clinical Psychology, 3, 1–27. 10.1146/annurev.clinpsy.3.022806.091432

Klein, B., Richards, J. C., & Austin, D. W. (2006). Efficacy of internet therapy for panic disorder. Journal of Behavior Therapy and Experimental Psychiatry, 37, 213–238.

Marks, I. M., Cavanagh, K., & Gega, L. (2007). Hands-on help: Computer-aided psychotherapy. Psychology Press.

Olthuis, J. V., Watt, M. C., Bailey, K., Hayden, J. A., & Stewart, S. H. (2016). Therapist-supported internet CBT for anxiety disorders in adults. Cochrane Database of Systematic Reviews, 3, CD011565. 10.1002/14651858.CD011565.pub2

Oromendia, P., Orrego, J., Bonillo, A., & Molinuevo, B. (2016). Internet-based self-help treatment for panic disorder: A randomized controlled trial comparing mandatory versus optional complementary psychological support. Cognitive Behaviour Therapy, 45(4), 270–286. 10.1080/16506073.2016.1163615

Orrego, J. (2025). Dataset for Panic Disorder Treatment: Clinical Efficacy and Anxiety Sensitivity Mechanisms in a Digital Intervention (Amindterapia / Start-Up Chile Project) [Data set]. Zenodo. 10.5281/zenodo.20084725

Sánchez-Meca, J., Rosa-Alcázar, A. I., Marín-Martínez, F., & Gómez-Conesa, A. (2010). Psychological treatment of panic disorder with or without agoraphobia: A meta-analysis. Clinical Psychology Review, 30(1), 37–50.

Sandín, B., & Valiente, R. (2007). ASI-3: Nueva escala para la evaluación de la sensibilidad a la ansiedad. Revista de Psicopatología y Psicología Clínica, 12, 91–104.

Santacana, M., Fullana, M. A., Bonillo, A., Morales, M., Montoro, M., Rosado, S., & Bulbena, A. (2014). Psychometric properties of the Spanish self-report version of the PDSS. Comprehensive Psychiatry, 55, 1467–1472.

Sanz, J., & Navarro, M. E. (2003). Propiedades psicométricas de una versión española del BAI. Ansiedad y Estrés, 9, 59–84.

Sanz, J., & Vázquez, C. (1998). Fiabilidad, validez y datos normativos del BDI. Psicothema, 10(2), 303–318.

Shear, M. K., Brown, T. A., Sholomskas, D. E., Barlow, D. H., Gorman, J. M., Woods, S. W., & Cloitre, M. (1992). Panic Disorder Severity Scale (PDSS). University of Pittsburgh School of Medicine.

Sheehan, D. V., Lecrubier, Y., Harnett-Sheehan, K., Janavs, J., Weiller, E., Bonora, L. I., & Dunbar, G. (1997). Reliability and validity of the MINI. European Psychiatry, 12, 232–241.

Sheehan, D. V., Harnett-Sheehan, K., & Raj, B. A. (1996). The measurement of disability. International Clinical Psychopharmacology, 11, 89–95.

Smits, J. A. J., Powers, M. B., Cho, Y., & Telch, M. J. (2004). Mechanism of change in cognitive-behavioral treatment of panic disorder: Evidence for the fear of fear mediational hypothesis. Journal of Consulting and Clinical Psychology, 72(4), 646–652. 10.1037/0022-006X.72.4.646

Ström, L., Pettersson, R., & Andersson, G. (2000). A controlled trial of self-help treatment via the Internet. Journal of Consulting and Clinical Psychology, 68, 722–727.

Taylor, S., Zvolensky, M. J., Cox, B. J., Deacon, B., Heimberg, R. G., Ledley, D. R., & Cardenas, S. J. (2007). Robust dimensions of anxiety sensitivity: Development and initial validation of the ASI-3. Psychological Assessment, 19, 176–188. 10.1037/1040-3590.19.2.176

